# Healthcare workers’ perceptions and attitudes towards the UK’s COVID-19 vaccination programme

**DOI:** 10.1101/2021.03.30.21254459

**Authors:** Louisa Manby, Anna Dowrick, Amelia Karia, Laura Maio, Caroline Buck, Georgina Singleton, Sasha Lewis-Jackson, Inayah Uddin, Samantha Vanderslott, Sam Martin, Cecilia Vindrola-Padros

## Abstract

**Objectives:** To explore healthcare workers’ (HCWs) perceptions and attitudes towards the COVID-19 vaccination programme in the UK, including their expectations, concerns and views on whether to promote vaccination to others. To understand the key factors shaping HCWs’ attitudes towards COVID-19 vaccination in the UK.

**Design:** This study was designed as a rapid qualitative appraisal integrating data from a review of UK policies and guidance on COVID-19 vaccination with data from in-depth semi-structured telephone interviews with frontline HCWs in the UK. Data were analysed using framework analysis.

**Participants:** Interviews were carried out with a purposive sample of HCWs from two large London-based hospital Trusts (n=24) and 24 government policies and guidelines on the vaccination programme were reviewed.

**Results:** The level of uncertainty about the vaccines’ long-term safety and efficacy against mutant strains made it difficult for HCWs to balance the benefits against the risks of vaccination. HCWs felt that government decisions on vaccine rollout had not been supported by evidence-based science and this impacted their level of trust and confidence in the programme. The spread of misinformation online also impacted HCWs’ attitudes towards vaccination, particularly among junior level and Black, Asian and Minority Ethnic (BAME) HCWs. Most HCWs felt encouraged to promote vaccination to their patients and the majority said they would advocate vaccination or engage in conversations about vaccination with others when relevant.

**Conclusion:** In order to improve HCWs’ trust and confidence in the UK’s COVID-19 vaccination programme, there needs to be clarity about what is known and not known about the vaccines and transparency around the evidence-base supporting government decisions on vaccine rollout. Effort is also needed to dispel the spread of vaccine-related misinformation online and to address specific concerns, particularly among BAME and junior level HCWs.

**Strengths and limitations of this study:**

- This is the first qualitative study to understand the factors influencing healthcare workers’ (HCWs) attitudes towards COVID-19 vaccination in the UK
- This study integrated interview and policy data and captured HCWs’ perceptions and attitudes in real-time as the vaccination programme was being rolled out in the UK
- Our interview study sample was limited in its representation of junior level HCWs and areas of the UK
- This research may have been impacted by selection bias as those with stronger views on vaccination may have been more likely to participate in the study

## Background

### COVID-19 and the UK’s COVID-19 vaccination programme

The coronavirus pandemic was declared a public health emergency on 11 March 2020 and, since then, has taken 2.61 million lives worldwide.^1^ In the UK, the government has implemented stringent measures, such as social distancing, the use of face masks and numerous national lockdowns in attempts to flatten the epidemic curve. Since December 2020, the UK has also been carrying out mass vaccination to provide the population with protection against COVID-19.^2^

The rollout of the mRNA Pfizer-BioNTech and viral vector Oxford-AstraZeneca vaccines reflects the largest vaccination programme in UK history.^3^ The UK’s vaccination programme is set to be rolled out from a mixture of hospitals, clinics, pharmacies and vaccination centres. A key element of the programme is the prioritisation scheme, developed by the Joint Committee on Vaccination (JCVI), which specifies who should receive the vaccine first. Frontline HCWs are listed as a priority group and the government has estimated that this will include 2.4 million HCWs in the UK.^2^ So far, crucial milestones have been met and by 15 February 2021, the government surpassed its target to vaccinate ∼14 million people in the first four categories of its prioritisation scheme.

Although the gains and cost-effectiveness of vaccines are undisputed, the individual and collective benefits achieved by them are ultimately contingent on the behaviours and attitudes of individuals.

## Known factors that influence attitudes towards vaccination

Vaccine hesitancy, which is defined by the World Health Organisation (WHO) as a ‘delay in acceptance or refusal of vaccines despite availability of vaccine services’, could be a potential barrier facing the COVID-19 immunisation programme.^4^ Vaccine hesitancy is a complex issue with many contributing factors, which vary across time, place and type of vaccine. ^4^ Hesitancy develops when there is a low perception of need for a vaccine, concerns over the efficacy and safety of the vaccine along with consideration of ease of accessing the vaccine. Determinants of vaccine hesitancy are complex and variable, fueled in part by misinformation, sociocultural factors, increasing individual empowerment and decreasing trust in governmental institutions.^5^ They are also highly context-specific e.g., non-uptake of the H1N1 vaccine in 2009 was related to the belief it had been expedited into circulation without rigorous testing.^6^

Data from the UK suggests that HCWs are perceived as the most reliable source of vaccine-related advice.^5^ A sense of duty in advocating vaccination to their patients, as well as belief in the benefits of immunisation programmes appear to be the primary reasons for HCWs to recommend uptake among patients.^5^ However, a number of obstacles can impede promotion, namely a lack of time to develop trusting relationships with patients, a lack of awareness of national vaccination guidelines, and personal reservations about the safety of certain vaccinations for specific populations.^7-9^ Most cited reasons for HCWs to self-vaccinate in non-pandemic contexts is to protect their patients, protect themselves and protect against absenteeism at work.^10^ Whilst numerous studies identify that HCWs are more likely to promote vaccination to patients if they have been vaccinated themselves^11^, research also demonstrates that self-vaccination rates among HCWs are in decline, particularly in relation to the Influenza vaccine.^11^ Concerns over safety and side effects, lack of time to get the vaccination for themselves, and belief they are at low risk are cited as the most prominent reasons for refusal.^12^ Vaccine hesitancy, from a patient or HCW perspective, does not necessarily result in vaccination refusal, but can cause delay in uptake and/or willingness to accept certain vaccines over others. ^13^

## HCWs’ attitudes towards the COVID-19 vaccination programme in the UK

Whilst there is much research relating to vaccine hesitancy among HCWs, the evidence base on vaccine hesitancy in relation to COVID-19, although rapidly emerging, remains limited. Given their increased risk of exposure to COVID-19 and high risk of transmitting the infection to vulnerable patients, it is crucial to protect HCWs from the virus through vaccination. There is a lot still unknown about the vaccines that have been developed to protect against COVID-19 and amidst this backdrop of uncertainty, HCWs are having to make sense of their feelings about COVID-19 vaccination and responsibilities within the COVID-19 vaccination programme.^14^ Recent research has identified that COVID-19 vaccine hesitancy exists among HCWs in the UK, particularly among certain groups.^15^ However, there is a lack of understanding about the factors shaping HCWs’ views on COVID-19 vaccination. Given that their intention to use and promote the vaccine to others is highly dependent on their knowledge and attitudes towards vaccination, it is crucial that we improve our understanding of these factors to guide public health communications encouraging vaccination among this group.^16^

## Methods

### Research questions

The principal research questions guiding this study were: 1) What are HCWs’ attitudes towards the vaccination programme? 2) What are the factors influencing HCWs’ attitudes towards vaccination? 3) What are HCWs’ attitudes towards promoting vaccination?

### Design

This study was designed as part of a larger, ongoing study investigating HCWs’ perceptions and experiences delivering care during the COVID-19 pandemic.^17^ This study utilised a rapid appraisal methodology with two different streams of data collection: telephone interviews with frontline HCWs and a review of UK government policies and guidance. Rapid appraisal designs often combine two or more different methods of data collection, which are then triangulated to improve the validity of the findings.^18^

### Sampling and participant recruitment

Interviews were conducted with a purposive sample of HCWs across two large Hospital Trusts in London. Purposive sampling was carried out to obtain a varied sample in terms of professional role, level of experience and ethnicity. Participants were approached by clinical leads at their Trusts who gathered verbal consent for the research team to contact them via email. Researchers then contacted participants and provided them with a participant information sheet and consent form. After receiving participants’ signed consent forms, researchers arranged a telephone interview via email at a convenient date and time. At the interview, participants were reminded that their participation was voluntary, they could withdraw from the study at any point, and researchers would ensure their anonymity was maintained.

The study was reviewed and approved by the Health Research Authority (HRA) (IRAS: 282069) and Research and Development (R&D) offices of the Trusts where the study took place.

### Data collection

Data collection and analysis methods are detailed in Table 1.

**Table 1:** Summary of data collection and analysis methods

| Type of data | Method of collection | Included sample | Method of analysis |
| --- | --- | --- | --- |
| Interviews | In-depth, semi-structured telephone interviews with frontline healthcare staff were conducted. | 24 interviews were conducted between 16 December 2020 and 3 March 2021. | Emerging findings were summarised using RAP sheets and verbatim transcripts were coded and analysed using framework analysis. |
| Policies | Relevant policies on the COVID-19 vaccination programme were selected from [legislation.gov.uk](https://www.legislation.gov.uk), https://www.england.nhs.uk and https://www.gov.uk/ | 24 policies and guidance documents published between 4 December 2020 and 15 February 2021 were identified. | Data were extracted into Excel by hand, cross-checked by another researcher and analysed using the analytical framework. |

### Interviews

In-depth, semi-structured telephone interviews were conducted with frontline HCWs. Interviews were carried out by researchers from the Rapid Research, Evaluation and Appraisal Lab (RREAL) using a semi-structured topic guide. Interview topics centred around HCWs’ perceptions towards the efficacy and safety of the COVID-19 vaccines as well as towards the delivery of the COVID-19 vaccination programme. All interviews were audio-recorded and transcribed verbatim. Researchers summarised emerging interview findings using rapid assessment procedures (RAP) sheets which improved familiarisation with the data and facilitated analysis whilst data collection was ongoing.^19^

### Policies

A review of policies and guidelines published by the UK government was conducted to contextualise HCWs’ experiences of the COVID-19 vaccination programme. Relevant policies were searched for using the search strategy detailed in Appendix 1. CVP selected policies that met the inclusion criteria (see Appendix 1), LM and SLJ then extracted and cross-checked the relevant data in Excel.

### Data analysis

All sources of data were analysed using the Framework Method guided by Gale et al. in order to triangulate the findings between the interview and policy data.^20^ RAP sheets were initially reviewed by all researchers for familiarisation purposes. After reviewing the RAP sheets and

interview transcripts, members of the research team developed an initial coding framework, which was guided by our principal research questions. In order to develop a timely analysis, we summarised our key findings using this coding framework set up in Excel. As the matrix allowed data from various sources to be compared, we were able to draw out the similarities and differences from the various data types for triangulation purposes.

## Results

In this section, we present the main emerging findings on HCWs’ attitudes and perceptions towards the COVID-19 vaccination programme (representative interview quotes are provided in Table 2).

**Table 2:** Representative interview quotes

| **Emerging findings** | **Representative quotes from interviews** |
| --- | --- |
| **Hopes for the COVID-19 vaccination programme** | *"We think it's going to end and it's not ending. So, the only real hope is a vaccine. I mean the only light at the end of the tunnel will be provided by mass vaccination; we're going to achieve mass immunity" (Surgeon)* |
| **Perceptions of vaccine effectiveness** | *"It does seem they will protect the population, particularly from the severer forms of COVID, which is where the problems arise" (Clinical director)* |
| **Perceptions of vaccine safety and risks** | *"I think we have to be hopeful that these vaccines, that they've been trialled and researched as much as they can to make sure we don't have any long-term effects." (Physiotherapist)*<br>*"So, we already know that the instance of adverse effects is very low. So, they seem safe and particularly if you balance the risk of having the vaccine against your risks of a serious illness with COVID, it's clearly in favour of having the vaccine" (Clinical director)* |
| **Degree of uncertainty about the COVID-19 vaccines** | *"Frankly, I'm scared to take the vaccine. I don't know if it's going to work, and I don't know if I want to put something in my body where there's no evidence to suggest that it is going to provide me with the immunity that I need to fight this virus" (General manager)* |
| **Government decisions on programme implementation** | *"Why the prime minister, who has no—clearly no medical competence, evidently, decides to change the scientific protocol with no collateral study to support this decision and no one agrees with him-- how is that possible?" (Nurse, sister)* |
| **Different sources of information on vaccination** | *"I have seen the people on higher grades, they went for the vaccine and they have had no issues but actually we have really, really struggled to engage people at the more junior level, and I think [social media] are having much more impact on them" (Divisional Manager)* |
|  | *"I was like, I'm not going to get the vaccine until I read something that reassures me that the overall scientific communities, and not just the UK say that it is safe to do" (Nurse, sister)* |
| **HCWs' attitudes towards promoting vaccination** | *"So, I guess it's, so, if it was a patient of mine or our teams and we really want, you know, we really advised them to have it done, I guess I would spend quite, myself or the nurse or the doctor, we would spend quite a lot of time understanding what their worries are, what their concerns are, and try and address that, and then, sort of, looking at the advantages and the disadvantages of having it or not having it. So, we would try and put the extra effort in to give them, it would be about them having a better-informed decision, rather than relying on things that they've read on social media or word of mouth from friends and family" (Clinical lead nurse)* |

### Theme 1: HCWs’ perceptions and attitudes towards the COVID-19 vaccination programme

#### Hopes for the COVID-19 vaccination programme

There was a general consensus among HCWs that vaccination offered a “*light at the end of the tunnel”*. They argued that vaccination was the only viable exit strategy, because they felt that infection prevention and control strategies which relied heavily on the behaviour of the general public had not been able to effectively control the spread of disease. Our policy review found that this was in line with the government’s view on the vaccination programme being a “foundation of our way out of this pandemic and the best way to protect people from COVID-19”.^21^ HCWs hoped that the vaccination programme would relieve pressure on the NHS by reducing the number, complexity and acuity of COVID-19 cases and that in doing so, would allow them to return to their usual standard of care. HCWs also noted the potential wider societal implications of vaccination, including its impact on the economy and on general day-to-day life. HCWs also commented that by reducing the transmission of the virus, vaccination could minimise the potential of the virus to mutate into more dangerous variants.

#### Perceptions of vaccine effectiveness

Our policy review indicated that the two-dose vaccine efficacy for the Oxford-AstraZeneca and Pfizer vaccines were 70.4% and 95% in phase III clinical trials respectively.^22^ HCWs felt that it was important to trust the scientific evidence on vaccine effectiveness, and they widely felt that the data available from clinical trials was convincing. In particular, HCWs were encouraged by the data that highlighted the vaccines were highly effective in reducing the severity of infections, given that the number of people with severe illness requiring hospitalisation and treatment in intensive care was reportedly the main factor causing strain on the healthcare system. Although HCWs were generally optimistic, some felt that the vaccines would not be totally effective in isolation. They argued that other infection prevention and control measures such as border controls would be necessary to reduce the global spread of infection whilst mass vaccination was rolled out worldwide. HCWs acknowledged the threat of mutant COVID-19 strains on the effectiveness of the vaccines, noting the difficulty of keeping the vaccines up to speed with changes to the virus. They feared this could compromise the vaccination programmes success. HCWs also noted that the success of the programme overall would rely on large-scale participation and political support.

#### Perceptions of vaccine safety and risks

Our policy review indicated that the frequency of severe adverse events following COVID-19 vaccination was low and that common side effects were short-lasting and included headaches, muscle aches and fever.^23^ Many HCWs felt that there was sufficient evidence available to demonstrate the short-term safety of the COVID-19 vaccines. HCWs commented that the known side effects were no different from those that arise from other vaccines. Concerns about vaccine safety generally pertained to the long-term, unknown side effects of the vaccines. In particular, there were concerns raised about the vaccines’ long-term impact on fertility. In balancing the risk of being vaccinated against the risk of experiencing severe illness from COVID-19, HCWs were generally prepared to accept the risk associated with vaccination to minimise their risk of severe disease from COVID-19.

### Theme 2: Factors influencing HCWs’ attitudes towards the vaccination programme

#### Degree of uncertainty about the COVID-19 vaccines

Hesitancy towards COVID-19 vaccination was driven, in part, by the lack of information available on the vaccines. Some HCWs who generally had confidence in vaccination and who reported to get the flu vaccine every year, expressed that they were unwilling to be vaccinated against COVID-19 due the lack of evidence available. There were also concerns that the development process for the vaccines was rushed as this meant a lot was still unknown about the vaccines when the programme was rolled out, which made it difficult to reach a decision. With new, emerging data and changing government guidelines, HCWs’ level of uncertainty increased, and they were left questioning *“what is evidence and what is not evidence”*.

There was a great deal of uncertainty among HCWs about the impact of mutant strains of the disease on vaccine effectiveness, as well as the vaccines’ optimum dosing schedules and long-term side effects. Given the amount unknown, some HCWs found it difficult to weigh up the risks and benefits of vaccination. Although some expressed that they wanted to wait for more evidence to emerge before making a decision, many ultimately decided that they were prepared to take the unknown risk given the lack of alternative options available. As the Oxford-AstraZeneca vaccine works by a known technology that has been tested previously with other vaccines, some HCWs expressed that they would be more confident to be vaccinated with this type over others.

#### Government decisions on programme implementation

The decision to prioritise vaccination for frontline health and social care workers and those at increased risk of vulnerability to infection was positively perceived.^24^ HCWs felt that it was important they were vaccinated to reduce sickness rates among staff and, ultimately, to ensure that the workforce available to treat patients was not compromised by infection with COVID-19. They also felt that it would help to reduce their levels of anxiety about becoming infected. Although the guidance was to prioritise among HCWs on the basis of their exposure and vulnerability to COVID-19, HCWs felt a sense of unfairness when this did not translate to practice as vaccines were predominantly given on a first-come-first-served basis.^24^

On 22 December 2020, a report was published by the government estimating that the efficacy of a single dose of either the Oxford-AstraZeneca or Pfizer vaccines was ∼73% and ∼90% respectively.^25^ Following on from this, the Department of Health and Social Care, with support from the JCVI, changed its recommendation to advise that the interval between vaccines doses be extended to 12 weeks.^26 27^ Widespread concern was expressed by HCWs about this decision. HCWs were worried that the decision was motivated by political agendas which ignored the evidence-based scientific protocols for the vaccines. Mixed and changing messages from the government were found to affect HCWs trust in the vaccination programme overall and there were concerns about there being *“no evidence or collateral study that says that it will be okay”*. HCWs felt that this went against the views of their Trusts and they called for more transparency from the government about the evidence supporting this. Very few HCWs felt that this was the right decision, and overriding feelings were of outrage and disappointment. These feelings were predominantly driven by fears that the decision would decrease their personal level of protection from COVID-19 and compromise the overall effectiveness of the vaccination programme. HCWs were also concerned that the delay between the doses would increase the likelihood of the second vaccine being missed due to people being *“lost in the ether of not knowing when they need to go back for their second dose”*.

#### Different sources of information on vaccination

Our policy review found several documents targeted at HCWs, addressing why it is important for them to be vaccinated against COVID-19 as well as providing information on the vaccines, their side effects and the level of protection they offer.^23 28^ Many HCWs reported that a key source of information they used to inform their decision on vaccination came from scientific communities and professional bodies. HCWs attitudes towards vaccination were also reported to be influenced by information being spread online, particularly that which was shared by senior clinicians and medical professionals. HCWs reported that the spread of misinformation on social media had ignited concerns and fears about the safety profile of the vaccines, particularly among certain groups. The impact of misinformation online appeared to vary depending on HCWs level of seniority and ethnicity, with more junior level and BAME HCWs being the most affected. There were reports about conspiracy theories being propagated online, many of which were targeted specifically at BAME groups with *“people saying that it is no coincidence that the majority of patients ventilated in ITU are Asian, it’s a deliberate genocide attempt and the vaccine has been engineered to adversely affect those populations. So, there is this kind of whole fake news circulating around the vaccine”*. Attitudes towards vaccination among HCWs’ family and friends were also noted to have an influence on their personal feelings towards the vaccines, particularly among BAME groups. Our policy review indicated that the government planned to take steps to involve BAME HCWs in conversations around vaccine rollout to ensure that their needs were understood and met.^21^

### Theme 3: HCWs’ attitudes towards promoting vaccination

Although HCWs reported that they did not feel obliged to promote vaccination, they felt strongly motivated to do so. Some HCWs believed they had a moral responsibility to advocate vaccination and they reported to play a very active role in encouraging their patients to get vaccinated by *“directing them, signposting them to links, phoning up patients who have questions about the vaccine*” with the aim of helping them to make informed decisions about vaccination. Many reported that patients often enquired whether they had been vaccinated. They felt that this was because patients wanted to ask HCWs that they trusted about their decision on vaccination for reassurance. However, not all HCWs advocated vaccination to patients and some felt that this did not fall under the remit of their role given that there was “*not much time and scope at the moment to discuss other things than the surgery they’re facing”*. Some HCWs also felt uncomfortable with the notion of trying to persuade people outside of their immediate family as they felt that it was everyone’s own right to make the decision for themselves; however, if directly asked by friends, colleagues or patients many said they would explain their reasoning behind being vaccinated.

## Discussion

During the rollout of the UK’s COVID-19 mass vaccination programme, HCWs had to rapidly make sense of their feelings about COVID-19 vaccination and responsibilities within the COVID-19 vaccination programme in a changing and uncertain context. The findings from this study demonstrate that, although HCWs were generally hopeful about the vaccines, their level of trust and confidence in the programme was impacted by political decisions and concerns about the unknown long-term side effects of the vaccines. HCWs were encouraged to promote vaccination and the majority of HCWs we interviewed said they would actively promote vaccination or engage in conversations about vaccination with others, where relevant.

Several other publications have also highlighted the impact of the unknown future effects of the COVID-19 vaccines on attitudes towards vaccination. ^14 29 30^ Paul et al., found that concerns about the vaccines’ unknown future effects led to feelings of ambivalence with almost one quarter (23%) of respondents saying that they were unsure about whether to be vaccinated against COVID-19.^14^ These publications also found that willingness to be vaccinated was impacted by mistrust in the safety of the COVID-19 vaccines. Generally, the level of confidence in government to handle the pandemic and to ensure the safety of the vaccines was low. ^14 29^ In our study, we found that concerns about vaccine safety were particularly prevalent among BAME and junior level HCWs and appeared to be fuelled, in part, by the spread of misinformation on social media. Several other publications have also revealed marked differences in attitudes towards COVID-19 vaccination among various sub-groups of the UK population. These studies have found associations between vaccine hesitancy and younger age, female gender, lower-income and ethnicity.^15 30 31^ Murphy et al., also found that respondents resistant to COVID-19 vaccination were less likely to obtain information about the pandemic from traditional and authoritative sources.^31^ Martin et al.’s observational analysis among HCWs demonstrated that differences in vaccine uptake were most marked in those of Black or certain South Asian HCW groups. This study also revealed that uptake varied depending on occupational group, with more administrative and executive staff (73.2%) reporting to be vaccinated compared with doctors (57.4%) and nurses (62.5%).^15^ This may reflect different opportunities among HCWs to get vaccinated, as we found in our study that vaccination was predominantly being delivered to HCWs on a first-come-first served basis.

We found that many HCWs advocated COVID-19 vaccination at both personal and professional levels. Verger et al., reported similar findings in their cross-sectional analysis undertaken among HCWs in France, Belgium and Canada in which 79.6% HCWs reported that they were likely to recommend the COVID-19 vaccine to their patients.^29^ Only 4.1% of HCWs in this study said that they were unlikely to promote vaccination and 16.3% were unsure. Interestingly, the proportion of HCWs who said they were likely to recommend the vaccine to their patients (79.6%) was higher than the proportion who said they were likely to be vaccinated themselves (72.4%), which indicates that some HCWs were likely to promote vaccination to their patients, even when they were personally hesitant about being vaccinated.

To our knowledge, this is the first qualitative study that has been undertaken to understand the factors influencing HCWs’ attitudes towards COVID-19 vaccination in the UK. This study captured HCWs’ perceptions and attitudes in real-time as the vaccination programme was being rolled out in the UK. As this study was conducted during the COVID-19 pandemic, telephone interviews were the most appropriate method for capturing HCWs’ perceptions and attitudes; however, this did make it more difficult for interviewers to pick up on participants’ non-verbal cues. Even though our policy review was national in scope, the participants we interviewed were from large, London-based hospital Trusts and this may impact the generalisability of our findings. Furthermore, although our sampling framework aimed to ensure a varied sample of HCWs, the majority of the HCWs we interviewed were senior level doctors and we had limited representation of junior level HCWs. Our research may have also been impacted by selection, bias as those with stronger views on vaccination may have been more likely to participate. As mass vaccination is still underway in the UK, our study will have missed any changes in attitudes since the end of the data collection period. It is important to recognise that attitudes to vaccination are constantly changing as both the pandemic and vaccination programmes progress. One multi-national meta-analysis of studies examining COVID-19 vaccination uptake found that the proportion of the population intending to vaccinate has declined as the pandemic has progressed.^32^

Firstly, our study highlights the importance of providing more clarity to HCWs on what is known and not known about the COVID-19 vaccines to enable them to feel more informed when making their decisions on vaccination. Secondly, our study also signifies the need for more clarity and transparency from government on the evidence basis for their decisions affecting vaccine rollout to improve HCWs’ confidence and trust in the vaccination programme overall. This study also highlights the need to directly engage with vaccine-hesitant sub-groups, including more junior level and BAME HCWs, understanding how a history of exclusion and racism might shape attitudes and practices in relation to vaccination^33^. NHS England has recently called for there to be one-to-one sessions arranged by line managers with vaccine hesitant HCWs to discuss the health benefits of vaccination. It is important that these conversations are handled carefully to enable open conversations to address HCWs’ concerns and dispel the mistruths that have been circulating online about the COVID-19 vaccines.^34^ There have also been recent reports that the government is considering making COVID-19 vaccination mandatory among frontline HCWs in the UK. In light of the findings of this study, it will be paramount that the UK government work to build trust and address uncertainty among HCWs before mandating vaccination in the UK.^35^

## Data Availability

All relevant data are included in the manuscript.

## Appendix 1 Policy review inclusion criteria

- Published between 1 December 2020 and 15 February 2021;
- Related to the COVID-19 pandemic;
- Related to the UK’s mass vaccination programme.

